# National Excess Mortality from Sub-National data: Method and Application for Argentina

**DOI:** 10.1101/2021.08.30.21262814

**Authors:** Ariel Karlinsky

## Abstract

Currently, many countries have not yet reported 2020 or 2021 mortality data to allow an estimate of excess mortality during the COVID-19 pandemic. However, some countries have sub-national mortality data, at the state, province or city level. I present a simple method to allow estimation of national level mortality and excess mortality from sub-national data, using the case of Argentina and projecting excess mortality up to August 2021.

## 1 Background

Excess mortality, the number of deaths above and beyond expected, is widely considered the gold-standard in quantifying the effects of pandemics such as COVID-19 (Beaney et al., 2020; Leon et al., 2020). Unfortunately, many countries have not yet reported 2020 or 2021 mortality data (Karlinsky and Kobak, 2021) thus inhibiting reliable estimates of excess mortality. One method to overcome this obstacle, is to project to countries without national level data using countries with national level data and the relation between the information in these countries and known properties of these countries, thus allowing out-of-sample prediction (The Economist, 2021; IHME, 2021). Another method, presented here, is to utilize reliable sub-national, i.e. state, province, city (from now on referred to as “local”), and projecting it to the national-level.

## 2 Estimation Method

To make things concrete, consider country *c*, that can be broken down to an *L* number of local units, each represented by *l*. The number of deaths *D* in the country, for every length of time, is thus a simple sum over the number of deaths in each local unit:

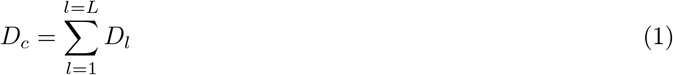

The share of deaths in the country represented by the local level deaths is thus:

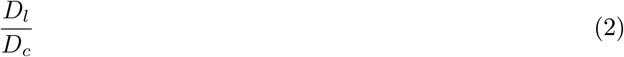

It follows that for every time period:

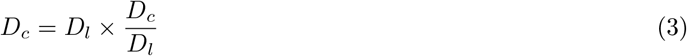

In order to use *D*_*l*_ to project to *D*_*c*_, we require an estimate for current 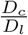, which is unknown since we do not have *D*_*c*_. We can estimate 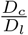 by using the historical ratio. This could be from before the pandemic or from a period during the pandemic where information on both the local mortality and the nation-wide mortality is available. This is more likely to hold where: (1) The pandemic’s spatial distribution is similar; (2) Movement between local units (migration) is low, so that state or province level local units are more suited than village, town or city level local units.

## 3 Application: Argentina

This method is applied to Argentina. Argentina is a federation of 24 provinces, which is the local unit. The province of Córdoba is the second largest province in terms of population size. *La Voz del Interior* is a Córdoban newspaper which has published monthly number of deaths in the province, ranging from January 2019 to August 2021 (Garbovetzky, 2021). This data was available a long time before Argentina has released 2020 mortality counts in Rearte et al. (2021), as collected in Karlinsky and Kobak (2021) and is much more recent, as latest national level data is only up to December 2020.

From 2005 to 2019, the share of annual deaths in Argentina accounted for by Córdoba was very stable, ranging from 8.55% to 8.84% (Ministry of Health, 2021b). In 2019, the ratio between the registered monthly number of deaths in Córdoba to that of all of Argentina ranged from 8.1% (January) to 10.5% (December), with a mean of 9% and the annual ratio being 9.0% as well.

In 2020, this monthly ratio ranged from 7.7% (August) to 10.3% (October) with a monthly mean of 9.0% as well (figure 1). The spatial distribution of COVID-19 deaths is very similar between Córdoba and the rest of Argentina (Ministry of Health, 2021a), as shown in figure 2. Thus, the projection of total mortality from Córdoba to the rest of Argentina during the COVID-19 pandemic is likely to hold.

**Figure 1:**
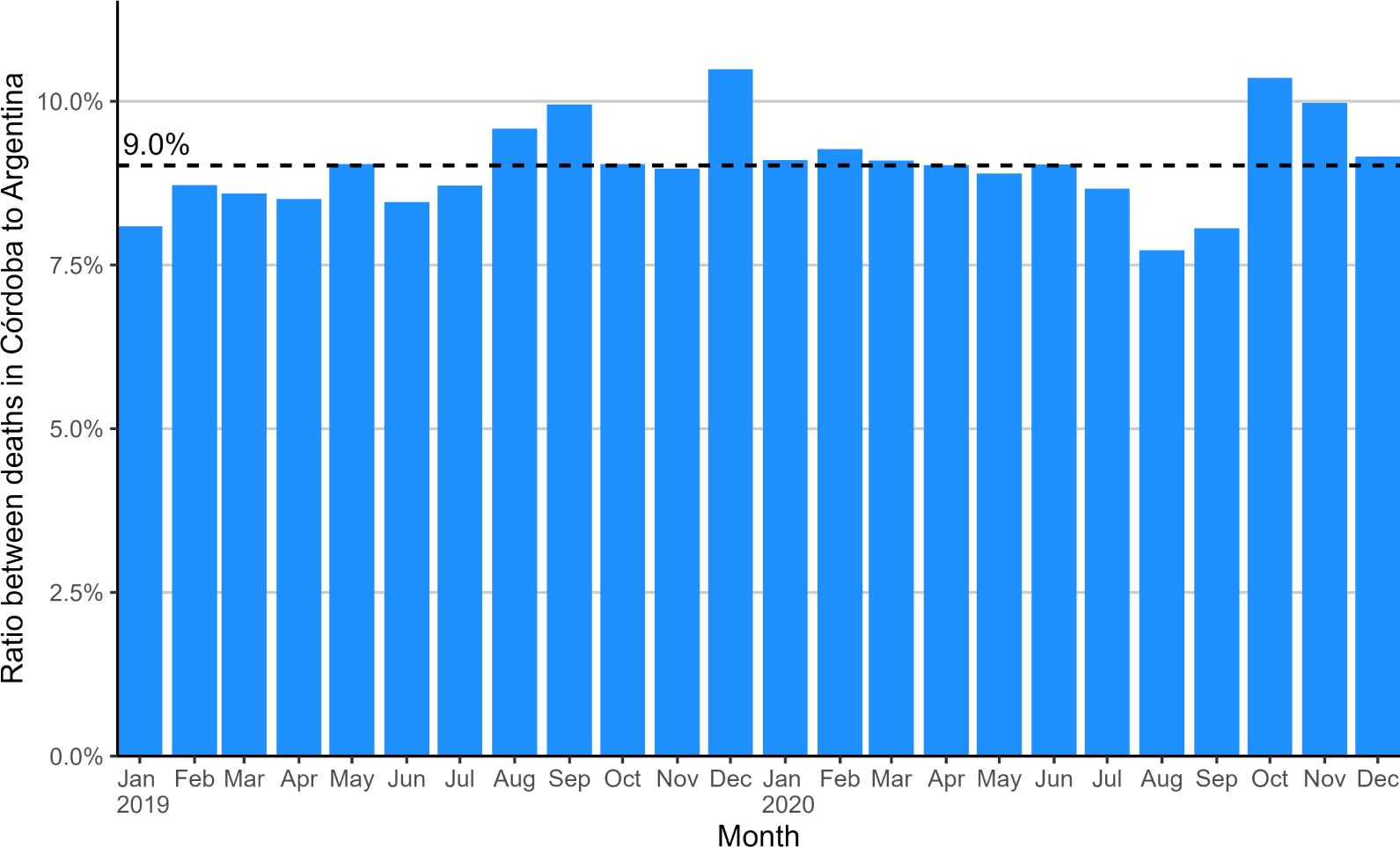
Ratio between deaths in Córdoba to total deaths in Argentina.

**Figure 2:**
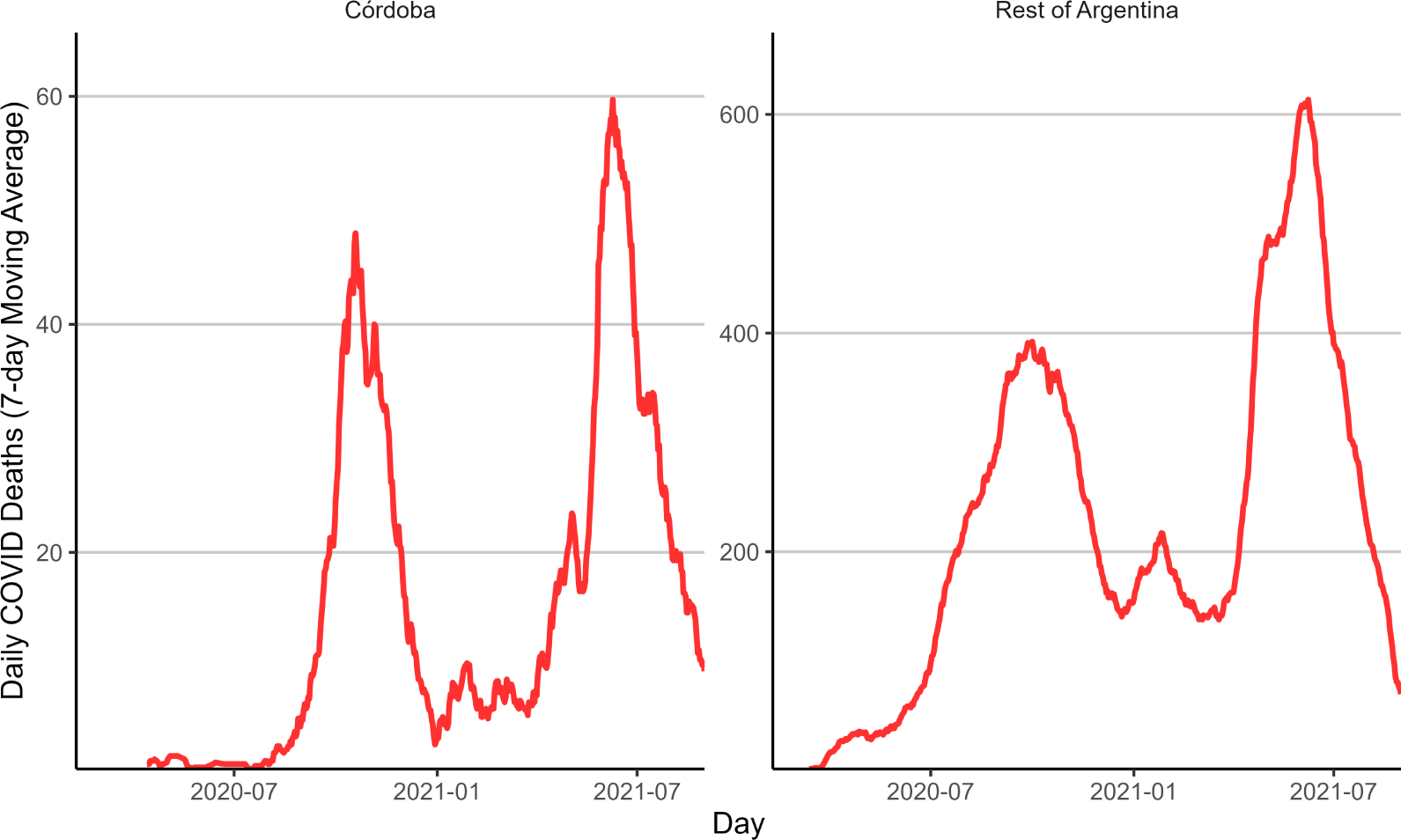
Daily COVID-19 deaths in Argentina in Córdoba and the rest of Argentina.

Using the reciprocal of 9.0%, to project monthly deaths from Córdoba to Argentina for 2021, monthly deaths are closely aligned between the registered national number and the projection in 2019 and most of 2020, as shown in figure 3. There are significant differences in August and September with total national mortality being higher, but this is then “compensated” by the projected mortality being higher in October and November, such that the total annual mortality is very similar, with 378,995 nationally registered deaths and 379,280 deaths as projected from Córdoba.

**Figure 3:**
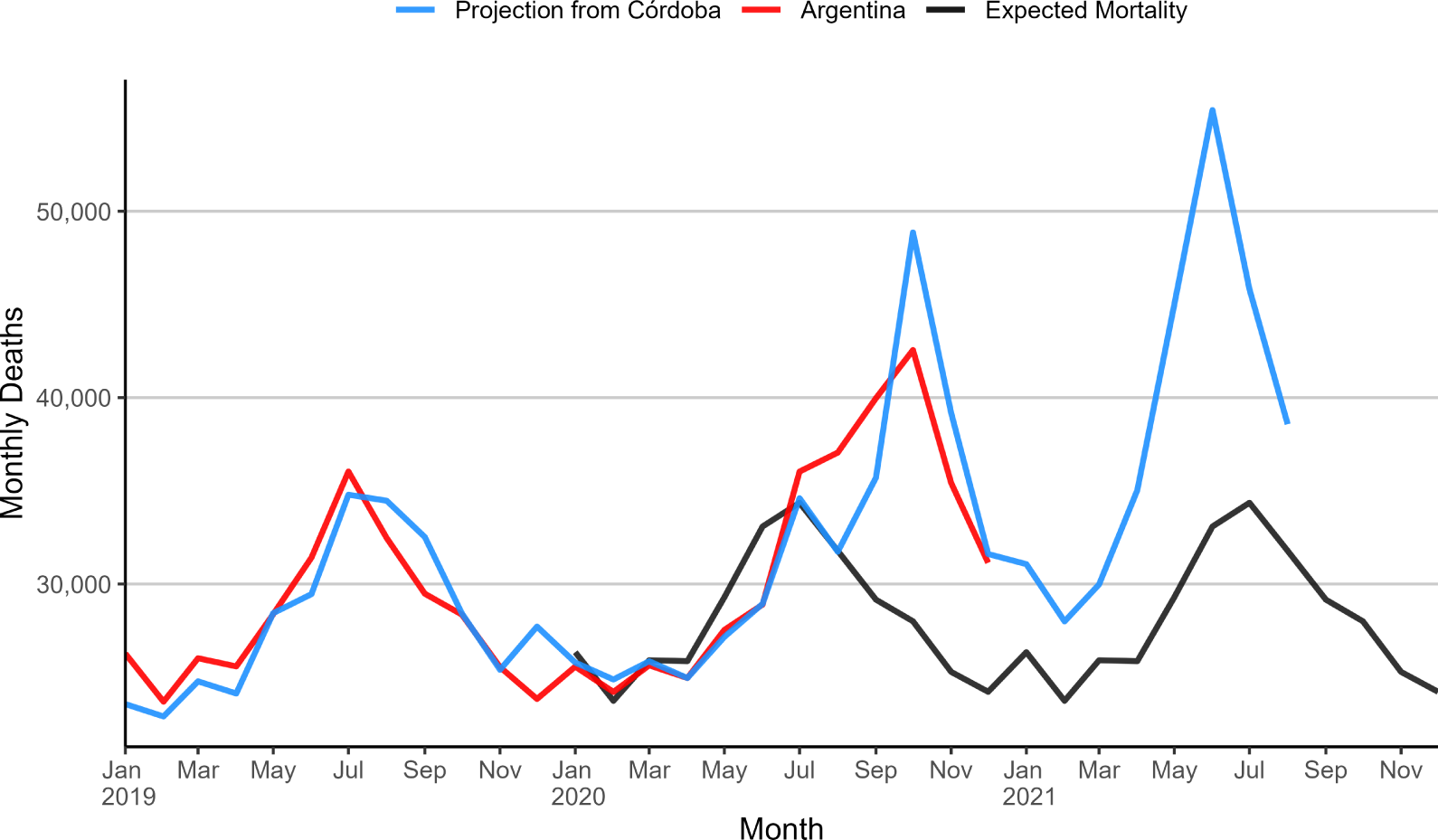
Monthly Deaths in Argentina.

Total excess from March 2020 to August 2021 (as computed by the Karlinsky and Kobak (2021) method) is 120,155 with COVID-19 deaths for the same period at 111,383 (WHO, 2021), amounting to an undercount ratio of 1.08 - similar to other Latin American countries like Brazil (1.09), Paraguay (1.06), Panama (1.05), higher than Chile (0.96), Peru (1.01) and lower than Bolivia (2.47), Ecuador (2.03) and Mexico (1.99) (Karlinsky and Kobak, 2021).

Similarly to other countries with fairly low undercount ratios, the cumulative (projected) excess deaths in Argentina tracks across time with the number of reported COVID-19 deaths (figure 4), lending further support to the projected excess mortality shown here.

**Figure 4:**
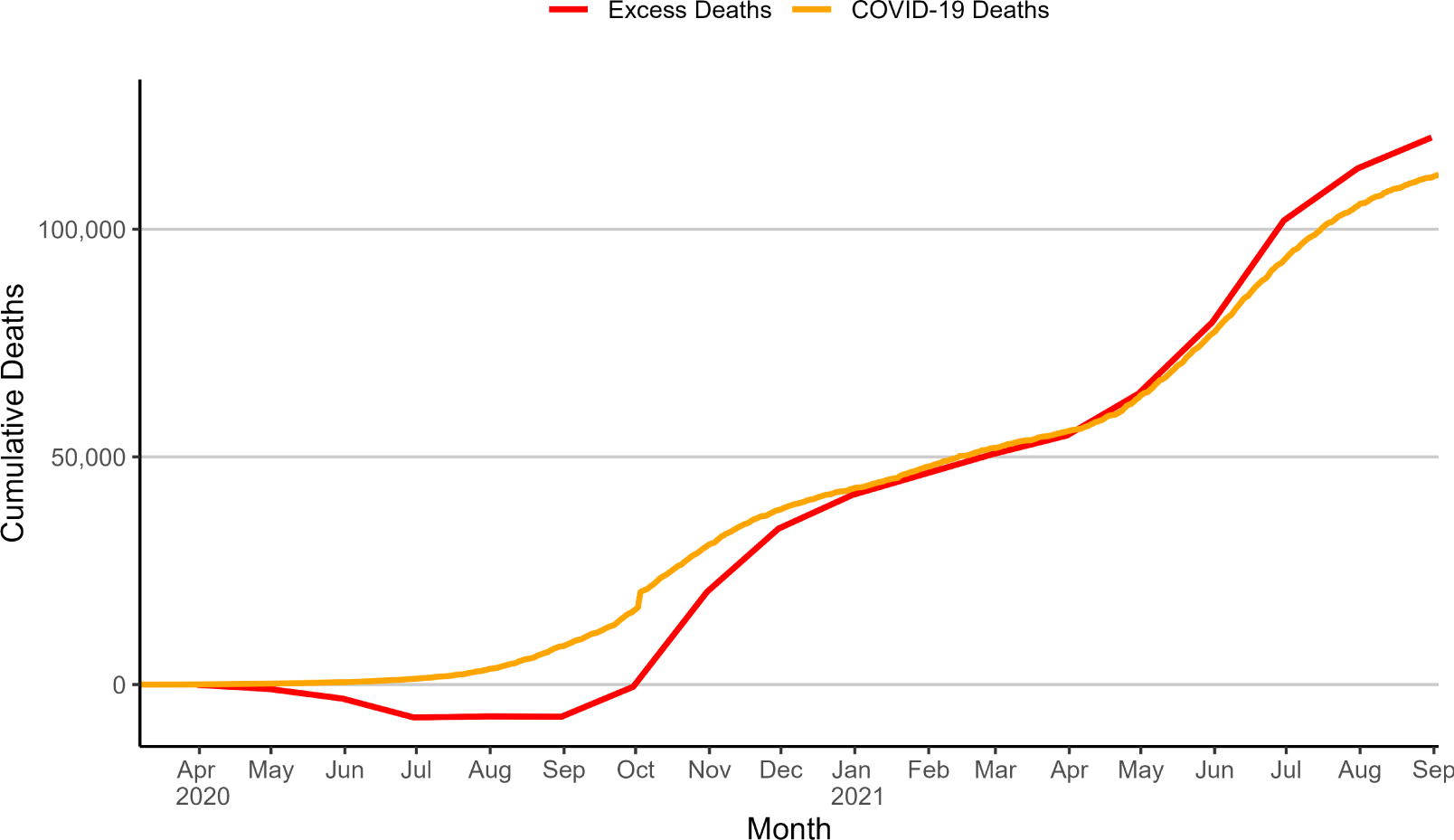
Tracking between reported COVID-19 deaths and Projected Excess Deaths in Argentina.

## 4 Discussion

The simple method I have shown in this paper allows derivation of national-level excess mortality during the COVID-19 pandemic, where only sub-national level data exists. The method is validated and shown to be plausible in the case of Argentina for 2020, and was used to estimate excess deaths up to August 2021. In several countries, sub-national data exists where national data has yet to be published. The method shown here can be applied to countries such as India (Leffler et al., 2021), Syria (Watson et al., 2021), Turkey (Yaman, 2021), Yemen (Besson et al., 2021), and more. It can also be utilized (as was shown here) in countries where national-level mortality data is more delayed than sub-national data, allowing almost real-time estimation of excess mortality.

## Data Availability

All data is publicly available at https://github.com/akarlinsky/world_mortality and the references cited.

